# Maternal gut microbiome-metabolome signatures implicate vitamin B12 insufficiency and one-carbon dysregulation in fetal spina bifida despite folate repletion

**DOI:** 10.64898/2026.11.21.26368660

**Authors:** Hauna Sheyholislami, Scott Dos Santos, Benjamin R. Joris, Vagisha Pruthi, Amanda J. MacFarlane, Gregory B. Gloor, Tim Van Mieghem, Kristin L. Connor

**Affiliations:** Department of Health Sciences, Carleton University; Sinai Health Systems, University of Toronto; Nutrition Research Division, Health Products and Food Branch, Health Canada; Department of Biochemistry, University of Western Ontario

**Keywords:** Pregnancy, neural tube defects, gut microbiome, circulating metabolome

## Abstract

Despite folic acid fortification, neural tube defects (NTDs), including spina bifida, persist, suggesting contributing factors beyond folate status. The maternal gut microbiome influences the availability of nutrients and metabolites essential for neural tube closure and fetal development, yet its role in NTDs and associated comorbidities remains unclear. We investigated and integrated data from maternal gut microbial and circulating metabolic profiles in pregnancies affected by fetal spina bifida (cases) and unaffected pregnancies (controls) using shotgun metagenomics and targeted metabolomics. Despite comparable folate status, cases had lower serum vitamin B12 and enriched microbial cobalamin biosynthesis genes, suggesting disrupted B12 metabolism. Lower serum vitamin E, elevated amino acids, and altered antioxidant pathways indicated altered oxidative stress-related metabolism. Reduced microbial pyrimidine degradation and circulating purine metabolism suggested disrupted nucleotide metabolism. These findings implicate vitamin B12 insufficiency, a pro-oxidative environment, and altered nucleotide metabolism as potential pathways contributing to pathophysiology of NTDs and their comorbidities.

## Introduction

Fetal neural tube defects (NTDs) occur in 1 in 2500 pregnancies in Canada^1^, and affect 244,000-322,000 pregnancies globally each year^2^. Two thirds of all NTDs are diagnosed as spina bifida, which is driving a recent rise in NTDs in the Canadian population^3^. NTDs result from unsuccessful closure of the neural tube during embryogenesis^4^, and are associated with additional comorbidities including placental maldevelopment^5^, intrauterine growth restriction^6^, lower body dysfunction, and developmental delay^7,8^. Folate and vitamin B12 are essential B-vitamins involved in nucleotide biosynthesis, homocysteine methylation, and cellular proliferation^9^; key processes for successful neural tube closure. Low circulating levels of maternal folate^10^ or vitamin B12^11^ increase the risk of NTDs including fetal spina bifida, which can be prevented with maternal periconceptional folic acid supplementation^12^ through a daily folic acid-containing multivitamin, and fortification of flour products^13^. Whilst these strategies prevent the incidence of NTDs, they are not completely adequate, as many women of childbearing age fail to reach recommended red blood cell folate levels to prevent fetal NTDs^14^ and many cases of fetal NTDs occur in women with normal folate blood concentrations^15^. This suggests that the mechanisms underlying folate and vitamin B12 transport and metabolism in pregnancy are not fully understood and there may be other unknown factors contributing to NTD risk.

The gut microbiome is a major contributor to host micronutrient levels. Specific gut microbes carry the genes for folate and vitamin B12 biosynthesis^16^ and are able to produce 30% of the daily recommended intake of these B vitamins^16^. Further, many gut microbes possess the biosynthesis pathways for purine and pyrimidine metabolism^17^, nucleotides that are needed for rapidly developing tissues and critical embryonic process, including neural tube closure^18^. Despite the involvement of the maternal gut microbiome in B vitamin and nucleotide metabolism, its potential role in NTD pathogenesis remains understudied, suggesting a gap in our understanding of mechanisms contributing to NTDs and their associated comorbidities, particularly in folate-replete populations.

Alterations in the gut microbiome may be reflected in the host’s circulating metabolome^19^, which includes the complete set of small molecule metabolites in the blood. These metabolites, including nutrients, amino acids, fatty acids, and lipids, among others, represent the byproducts of both host and microbial metabolism^20^, reflect dynamic biological and metabolic processes in the host, and in pregnancy, could therefore influence embryonic/fetal development^21,22^. Previous studies have described select maternal circulating metabolite levels in the context of NTDs, but have not considered the role of the maternal gut microbiome and the circulating metabolome together, which may uncover previously unknown mechanisms involved in NTD pathogenesis and poor fetal outcomes in these pregnancies.

To determine whether and how alterations in the maternal gut microbiome and circulating metabolome reflect metabolic disruptions relevant to NTD and comorbidity risk beyond folate status, we conducted a matched case-control study comparing pregnancies carrying a fetus with spina bifida (cases) and unaffected pregnancies (controls). We first characterised differences in maternal gut microbial composition and function and in circulating nutrient and metabolite profiles between case and control pregnancies, then integrated these datasets to determine whether gut microbial alterations were associated with systemic metabolic differences. We additionally estimated gut microbial biosynthetic capacity for folate, vitamin B12, and purines and pyrimidines to determine whether differences in this capacity might contribute to case metabolic signatures. We hypothesised that case mothers would have an altered gut microbiome, characterised by differences B vitamin and nucleotide metabolic functions, reflected in altered levels of circulating metabolites and metabolic pathways important for neural tube closure and fetal growth. We also hypothesised gut microbial biosynthetic capacity for these substrates would differ between groups.

## Methods

This study was approved by the Carleton University Research Ethics Board (108884), the Mount Sinai Hospital Research Ethics Board (17-0028-E) and the Health Canada Research Ethics Board (2025-031H). All participants provided informed consent upon enrolment.

### Participants

In this case-control study, 40 pregnant people were recruited from Mount Sinai Hospital (MSH) in Toronto, Ontario, Canada. Pregnant people carrying a fetus with spina bifida (cases) were recruited from the fetal medicine unit at MSH. Pregnant people carrying a fetus without spina bifida (controls) were recruited from the low-risk obstetric unit at MSH. Cases and controls were recruited at ∼25 weeks gestational age. Pregnancies that were associated with other structural or genetic fetal anomalies were excluded from both groups.

### Sample collection and study procedures

Maternal medical history was obtained including age, ethnicity, height, weight, body mass index (BMI) before pregnancy, pre-pregnancy diabetes, a previous pregnancy with NTDs, family history of NTDs and fetal anomalies, abortus, gravidity, parity, gestational age at recruitment, pregnancy complications, other fetal anomalies, dietary restrictions, daily probiotic supplementation across gestation and prenatal supplementation. Maternal rectal swabs were collected at approximately 25 weeks’ gestation for gut microbiome composition, along with a maternal blood draw for serum metabolome characterization. A one-time standardized 24-hour dietary recall questionnaire was administered at recruitment using the Automated Self-Administered 24-hour Recall (ASA24™) for Canada to estimate macronutrient and micronutrient intake.

### Metagenomic sequencing and analysis

The full methods and references used for metagenomic sequencing and analyses are detailed in Supplemental Methods. In brief, total genomic DNA was extracted from rectal swabs of cases (n=17) and controls (n=17) using the QiAmp PowerFecal Pro DNA Kit and sequenced on an Illumina NovaSeq (75 nt paired-end reads; London Regional Genomics Centre, London, ON). Reads aligning to the human genome (GRCh38 and T2T-CHM13v2.0) were removed using Bowtie2, and the remaining non-human reads were de-duplicated (BBMap), quality-assessed (FastQC, MultiQC), and trimmed (Trimmomatic). Metagenomes were assembled with metaSPAdes and MEGAHIT within MetaWRAP, human-classified contigs were removed (Kraken2), and contigs were binned with MetaBAT2 and MaxBin2, refined, and reassembled, retaining bins (i.e. taxa) that were >75% complete with <10% contamination. Bins were dereplicated at 99% similarity (dRep), taxonomically classified with the GTDB toolkit (release 220), and quantified by mapping reads back to bins (Bowtie2, samtools). For functional profiling, genes were called with Prodigal in an Anvi’o contig database, KEGG orthology (KO) terms were assigned by HMM search against Kofams, and each KO term was manually curated to a single KEGG pathway. Reads were mapped to the called genes and aggregated by KO term to generate a functional feature table. Read count tables for taxa and functions were batch-corrected with ComBat_seq (sva) and analysed for differential abundance with ALDEx2 using scale simulation, with effect size cutoffs of >0.5 for taxa and >0.35 for functions (corresponding to a medium Cohen’s d), where positive values indicate overrepresentation in cases and negative values overrepresentation in controls.

### Functional exploratory analysis

The metabolic capability of each dereplicated bin was estimated using metabolic pathway information provided by KEGG, through the ‘anvi-estimate-metabolism’^23^ function of the Anvi’o package. The output provided was a list of KEGG pathways that were identified in each dereplicated bin. To determine high-level differences in metabolic capabilities of differentially abundant taxa (i.e. bins) in cases and controls, the KEGG pathways identified were extracted for the overrepresented taxa in cases and controls (d>0.5). Given that these taxa are overrepresented in cases and controls, it is plausible that their metabolic capabilities may also have a biological impact. The KEGG pathways identified in the overrepresented taxa were then counted and grouped to determine the total number of each pathway within overrepresented taxa in cases and controls separately. Using the final pathway counts, pathways related to NTDs (including vitamin and cofactor metabolism and DNA synthesis) were extracted. Fisher’s exact tests were conducted on all NTD-related pathways to determine differences between cases and controls (FDR q<0.05).

### Targeted metabolomics of serum samples

Serum samples (500ml/participant; 20 cases and 20 controls) were analyzed using four targeted quantitative metabolomics assays (The Metabolomics Innovation Centre [TMIC], University of Alberta): fat-soluble vitamins (vitamins A, D and E; UPLC-MRM/MS);, water-soluble vitamins (vitamins C, B7, B3, B3-amide, B6, B2 and B1; UPLC-MRM/MS); metals/minerals (41 metals and minerals; ICP-MS); and TMIC MEGA Clinical Assay 2.0 (600 metabolites, including amino acids, lipids, fatty acids, bile acids; UPLC-MRM/MS). Serum folate concentrations were measured using the Centers for Disease Control and Prevention (CDC) folate microbiological assay and vitamin B12 concentrations were measured using the Siemens Atellica clinical analyser equipped with the vitamin B12 reagent packs, as per manufacturer’s protocols (Ottawa, Ontario, Canada).

### Serum metabolomic analyses

All metabolomic analyses were performed on the MetaboAnalyst platform (v.6.0). First, differentially abundant metabolites between cases and controls were identified using Student’s t-tests with normalized (log2 transformed) concentrations (FDR q<0.05). Metabolites below the limit of detection were excluded. Second, metabolite set enrichment analysis (MSEA) was used to identify up- and downregulated metabolite sets (pre-determined by MetaboAnalyst based on similar functions). Third, pathway topological analysis was used to identify significantly impacted metabolic pathways, with pathway impact calculated from relative-betweenness centrality (higher scores reflecting more central positions of altered positions within a pathway). For both MSEA and pathway analysis, up- or downregulation was defined by the direction of change in the underlying metabolites within the metabolite set/pathway (FDR q<0.05).

Metabotyping analysis was also conducted to identify metabolomic profiles that may discriminate samples^24,25^. Normalized metabolite concentrations of all MEGA metabolites (e.g. amino acids, lipids, triglycerides, etc.) were used for hierarchal clustering, where the silhouette method was used to determine the optimal number of metabotypes (k means=2)^26^, which evaluates how well-matched each metabolite is to its assigned cluster relative to other clusters^26^.

### Exploratory analysis of microbial production capabilities of folate, vitamin B12 and DNA nucleotides

To explore the potential capacity of the maternal gut microbiome to produce folate, vitamin B12, and purines and pyrimidines, and whether this differed between cases and controls, we conducted an analysis using KEGG modules. Full methods are detailed in Supplemental Methods. In brief, folate^27^ production (tetrahydrofolate biosynthesis pathway; M00126, tetrahydrofolate biosynthesis pathway, mediated by PTPS; M00841), vitamin B12^27,28^ production (anaerobic cobyrinate biosynthesis pathway; M00924, aerobic cobyrinate biosynthesis pathway; M00925, cobalamin biosynthesis pathway; M00122), and purine/pyrimidine biosynthesis (purine synthesis; M00048, pyrimidine synthesis; M00051, thymidylate synthesis; M00938) and degradation (adenine degradation; M00958, guanine degradation; M00959, pyrimidine degradation; M00046, alternative pyrimidine degradation; M00939) were assessed using pathwise module completeness scores (defined sets of enzymatic steps in a pathway; scored 0-1)^23^ in each taxa. For vitamin B12, the higher upstream score (e.g. the aerobic/anaerobic pathways producing cobyrinate a,c diamide; M00924/M00925) was compared with M00122 (the downstream pathway producing cobalamin from cobyrinate a,c-diamide), retaining the lower (rate-limiting) value. Completeness scores were weighted by taxa relative abundance and summed across taxa to yield individual (participant)-level capacity scores. Mean capacity scores between cases and controls were compared using Student’s t-test (FDR q<0.05).

### Dietary recall analysis

In brief, dietary intake was assessed using the Automated Self-Administered 24-hour Recall (ASA24-Canada-2016 & 2018), a Canadian adaptation of the National Institute of Health’s ASA24 developed with Health Canada, incorporating nutrient data from the Canadian Nutrient File. Dietary recall questionnaires were administered at recruitment and completed by 18 participants in each group (n=36), with outlier entries (total energy intakes <600 or >4400 kcal/day) excluded per ASA24 guidance^29^. Healthy Eating Index (HEI)^30^ scores were calculated from reported food intake to generate adequacy component scores (total and whole fruits, total vegetables, greens and beans, whole grains, dairy, total protein foods, seafood and plant proteins, fatty acids), and moderation component scores (refined grains, sodium, added sugars, saturated fats), summed into a total HEI score and classified as good (>80), needs improvement (51-80), or poor (<51). Dietary Inflammatory Index (DII)^31^, based on 27 nutrients and two food items (onions and garlic), was also calculated to estimate dietary inflammatory potential. Scores ranged from -8.87 to 7.98, where negative scores indicate a diet with anti-inflammatory potential, 0 indicates a neutral diet, and positive scores indicate a diet with pro-inflammatory potential. Full methods are included in Supplemental Methods.

### Statistical analysis of baseline participant characteristics

An *a priori* sample size calculation was performed based on a previous study of the gut microbiome during pregnancy^32^. A minimum sample size of 16 cases and 16 matched controls was estimated to provide 80% statistical power to detect a 30% between-group difference in gut metagenomic outcomes, at a significance level of 0.05. To account for potential loss to follow up, the recruitment target was increased to 20 cases and 20 matched controls. The final sample included 17 cases and 17 matched controls (n=34) for gut metagenomic analyses, and 20 cases and 20 matched controls (n=40) for serum metabolomic analyses. Maternal clinical, demographic and dietary intake data were analyzed using JMP (Student edition 18; SAS Institute). Differences between cases and controls for continuous maternal clinical and demographic outcomes, intake of individual macro- and micronutrients, and HEI and DII scores were analysed using student’s t-tests (parametric data) and the Wilcoxon Rank Sum test (non-parametric). Categorical maternal clinical and demographic outcomes were analysed using Likelihood Ratio Chi Square test. Continuous data are reported as median (interquartile range) with p/q values from t-test/Wilcoxon, and categorical data are n (%) with p values from Likelihood Ratio Chi Square. Significance was set at p/q<0.05.

## Results

### Study characteristics

Socio-demographic characteristics of women who had a fetus with spina bifida (n=20) and women with an unaffected fetus (n=20) are summarized in Table 1. As expected from the matched study design, case and control groups had similar in BMI and race/ethnicity (Table 1). There were differences in maternal age and fetal birthweight between groups (p<0.05). Most women in both groups identified as White, and most women reported use of prenatal vitamins (Table 1).

**Table 1.** Participant clinical and demographic characteristics.

|  | Controls (n=20) | Cases (n=20) | p value |
|---|---|---|---|
| Maternal age (years) | 34.6 (33, 36) | 30 (28, 31) | 0.0005* |
| Maternal race/ethnicity (n [%]) |  |  |  |
| Asian | 2 (10) | 2 (10) | 1 |
| Black | 0 | 0 |  |
| Latin/Hispanic | 0 | 0 |  |
| White | 18 (90) | 18 (90) |  |
| Other | 0 | 0 |  |
| Pre-pregnancy BMI (kg/m<sup>2</sup>) | 23.8 (21.7, 28.6) | 24.6 (21.8, 28.7) | 0.66 |
| Pre-pregnancy BMI classified (n [%]) |  |  |  |
| Underweight (<18.5) | 1 (5) | 1 (5) | 1 |
| Normal weight (18.5-24.9) | 12 (60) | 12 (60) |  |
| Overweight (25-29.9) | 3 (15) | 3 (15) |  |
| Obese (>30) | 4 (20) | 4 (20) |  |
| Fetal birthweight | 3.32 (2.89, 3.61) | 2.46 (1.6, 3.03) | 0.0003* |
| Prenatal vitamins (n [%]) | 15 (75) | 16 (80) | 0.70 |
| Probiotics (n [%]) | 5 (25) | 3 (15) | 0.43 |

Data are median (IQR; Student’s t-test for normally distributed data with equal variance or Wilcoxon test for non-parametric data or data with outliers) for continuous variables or n (%) (Likelihood Ratio Chi Square test) for categorical variables. \*p < 0.05. BMI = body mass index. IQR = interquartile range.

### Cases exhibit vitamin B12 insufficiency, reduced antioxidant nutrient status, and a pro-inflammatory dietary profile, despite comparable folate status

Serum levels and dietary intake of folate and vitamin B6 were equivalent in cases and controls, but cases had lower dietary intake (p=0.03) and serum levels (p=0.02) of vitamin B12, compared to controls (Table 2; Figure 1). Dietary intakes of vitamin C and choline (p=0.005 and p=0.03, respectively; Table 2), and serum vitamin E (alpha-tocopherol) levels (q=0.005; Figure 1), were also significantly lower in cases compared to controls. HEI adequacy scores were not different between groups, but cases had lower HEI moderation scores for refined grains, suggesting higher dietary intake (p=0.0002; Table 3). Whilst DII scores did not differ significantly between groups, cases had a positive total DII score (0.84 [-0.15, 1.60]; Table 3), compared to a neutral score for controls (-0.08 [-0.81, 0.87]), suggesting a more pro-inflammatory diet.

**Table 2.**
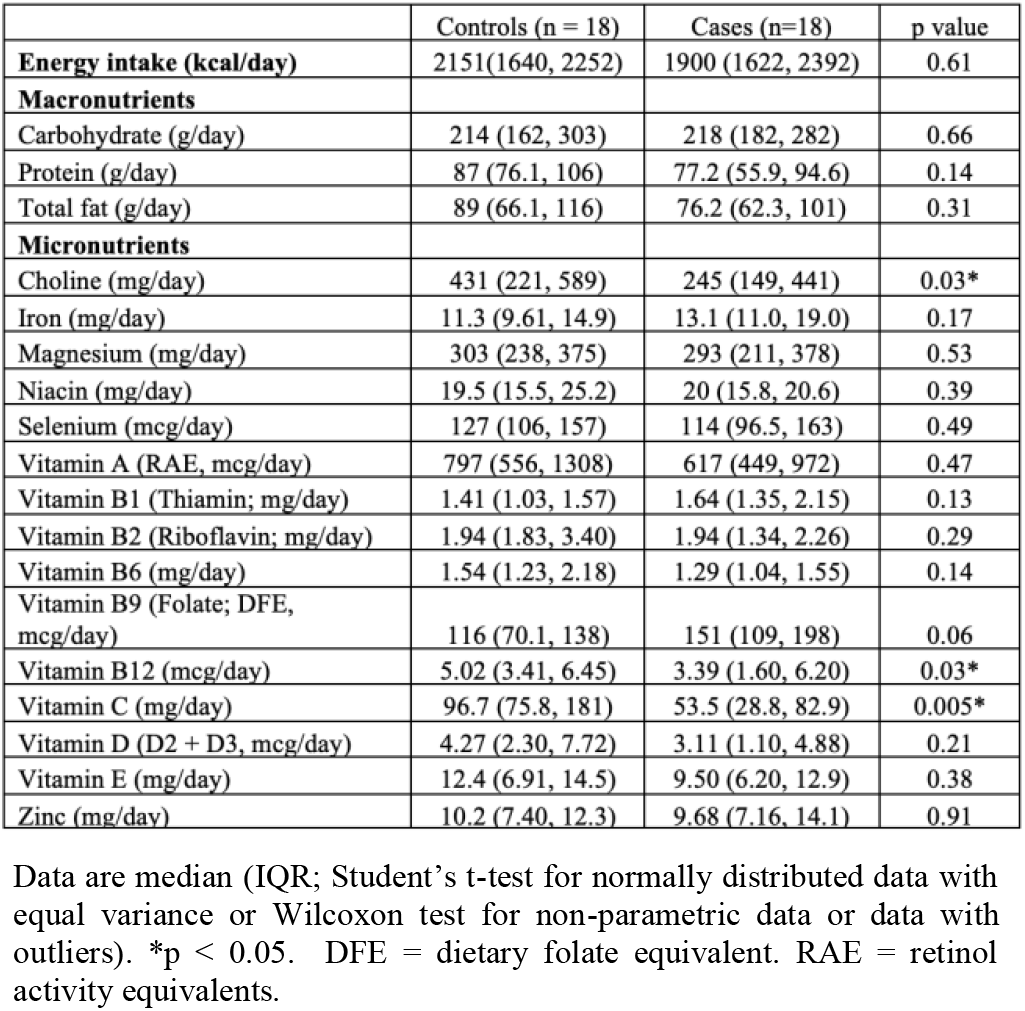
Self-reported dietary intake of macronutrients and micronutrients.

**Table 3.** Maternal Healthy Eating Index (HEI) and Dietary Inflammatory Index (DII) scores derived from dietary recall data.

| Dietary Components | Score range | Controls (n=18) | Cases (n=18) | p value |
|---|---|---|---|---|
| **HEI components** |  | Median (IQR) | Median (IQR) |  |
| *Adequacy score* | 0-60 | 34.6 (28.2, 38.8) | 32.7 (22, 38.7) | 0.23 |
| Total fruits | 0-5 | 2.68 (0.61, 5) | 1.03 (0.44, 3.54) | 0.24 |
| Whole fruits | 0-5 | 4.10 (0.49, 5) | 1.28 (0.26, 5) | 0.18 |
| Total vegetables | 0-5 | 4.63 (2.1, 5) | 3.50 (2.53, 4.40) | 0.34 |
| Greens and beans | 0-5 | 1.92 (0, 5) | 2.94 (0, 5) | 0.82 |
| Whole grains | 0-10 | 0.12 (0, 2.31) | 1.43 (0, 4.77) | 0.48 |
| Dairy | 0-10 | 7.86 (3.08, 10) | 6.26 (4.54, 8.45) | 0.49 |
| Total protein foods | 0-5 | 5 (3.53, 5) | 5 (2.75, 5) | 0.83 |
| Seafood and plant proteins | 0-5 | 5 (0.27, 5) | 1.59 (0, 5) | 0.15 |
| Fatty acids | 0-10 | 5.23 (3.16, 9.67) | 4.01 (2.11, 7.82) | 0.34 |
| *Moderation score* | 0-40 | 21.9 (16.6, 26.4) | 18.7 (16.7, 20.1) | 0.19 |
| Refined grains | 0-10 | 7.90 (6.37, 10) | 2.32 (0, 6.82) | 0.0002* |
| Added sugars | 0-10 | 8.44 (6.02, 10) | 8.97 (6.35, 9.53) | 0.91 |
| Sodium | 0-10 | 1.49 (0, 3.35) | 2.60 (0, 5.55) | 0.45 |
| Saturated fats | 0-10 | 4.14 (0.17, 6.29) | 6.04 (1.57, 8.30) | 0.27 |
| *Total score* | 0-100 | 54.3 (48.2, 63.6) | 49.0 (42.9, 55.1) | 0.09 |
| HEI score rating (n [%]) |  |  |  | 0.59 |
| Good (>80) |  | 1 (5.6) | 1 (5.6) |  |
| Needs improvement (51-80) |  | 10 (55.6) | 7 (38.9) |  |
| Poor (<51) |  | 7 (38.9) | 10 (55.6) |  |
| **Dietary inflammatory index** |  |  |  |  |
| Total DII score |  | -0.08 (-0.81, 0.87) | 0.84 (-0.15, 1.60) | 0.1 |

Data are median (IQR; Student’s t-test for normally distributed data with equal variance or Wilcoxon test for non-parametric data or data with outliers). \*p < 0.05. HEI = Healthy Eating Index. DII = Dietary Inflammatory Index.

**Figure 1.**
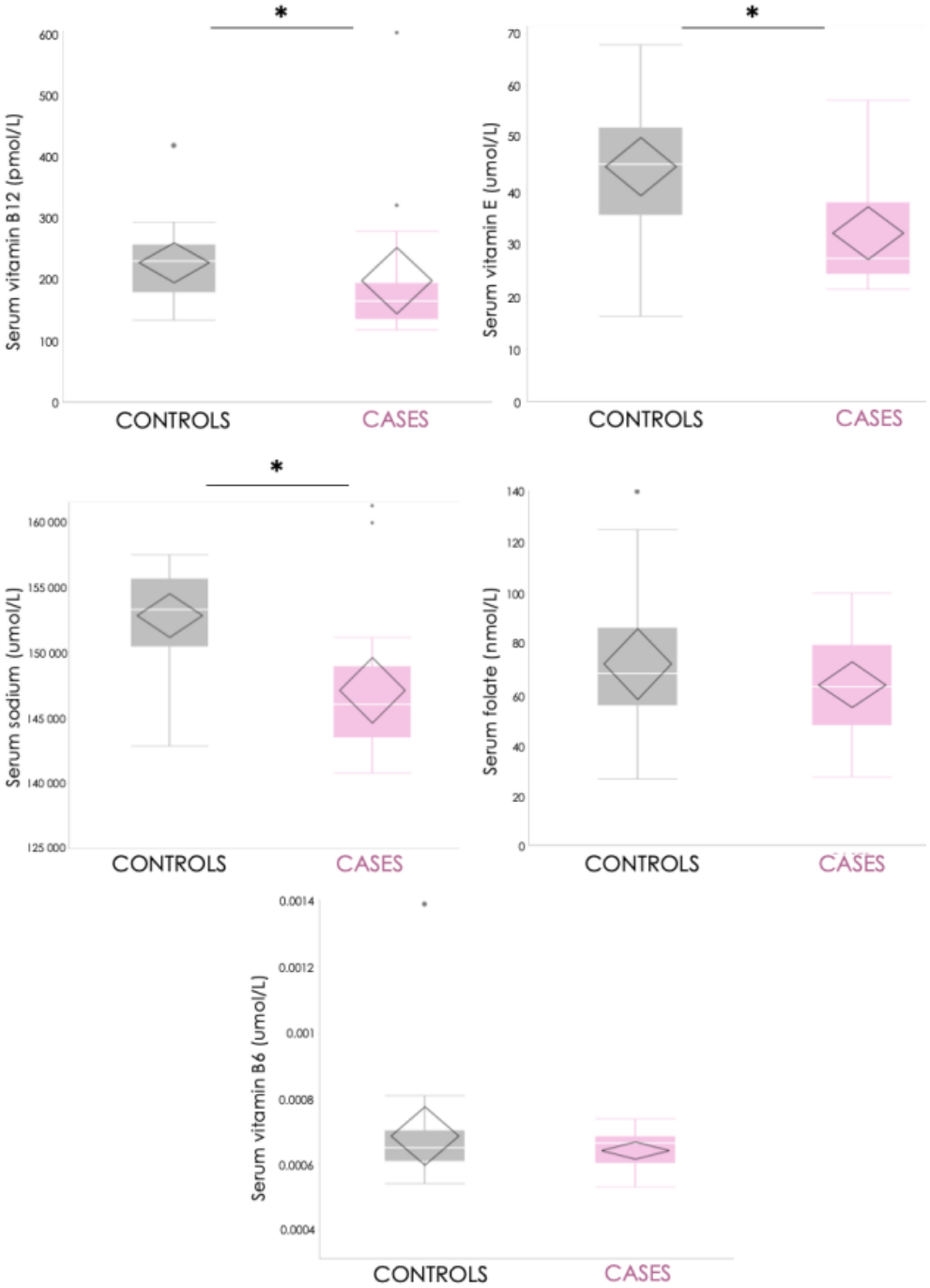
Serum vitamin B12, vitamin E, sodium, folate and vitamin B6 levels in cases and controls. Data are quartiles (median lines) and whiskers extend to 1.5 x interquartile range. Points outside this range are plotted as outliers. Data were compared between groups using a two-tailed Student’s t-test of log-transformed serum nutrient concentrations (FDR-corrected) or Wilcoxon rank sum test for data with outliers. *p/q < 0.05, statistically significant.

### Cases harbour gut taxa linked to adverse pregnancy outcomes and enriched gut microbial lipid, amino acid and cobalamin functions

To first determine whether and how composition of the maternal gut microbiome in cases differed compared to controls, differential abundance analysis of taxonomic assignments was performed. Twenty-two bacterial taxa were overrepresented in cases compared to controls (d*>*0.5; Figure 2A). These taxa belonged to the genera *Fenollaria* (n=5), *Porphyromonas* (n=4), *Prevotella* (n=3) *Agathobacter* (n=2), *Corynebacterium* (n=2), *Murdochiella* (n=1), and *Bifidobacterium* (n=1) genera (Figure 2A). The remaining taxonomic assignments did not have an identified genera due to lack of identification of these bacteria in the reference database (unnamed genus belonging to the order Saccharofermentales (SFGY01; n=4). The most overrepresented taxon in cases was *Fenollaria timonensis* (d=0.72; previously identified in cord blood of stillborn infants and those born preterm^33^), followed by an unidentified taxon from the *Porphyromonas* genus (species ID: sp900546675; d=0.68) of gram-negative, anaerobic bacteria (Figure 2A). The most studied species of this genus, *P. gingivalis*, is a pathogen responsible for periodontal disease, and has been implicated in pathophysiology of preterm birth, preeclampsia^34^ and fetal growth restriction^35^ in pregnancy. Conversely, only two bacterial taxa were overrepresented in controls compared to cases: *Streptococcus vaginalis* (d=-0.56) and *Escherichia coli* (d=-0.58; Figure 2A). We next aimed to determine differences in functional capabilities of the maternal gut microbiome between cases and controls. Twenty-two biological pathways were overrepresented in cases compared to controls (Figure 2B). These included six functions within the porphyrin metabolism pathway; KO terms corresponding to CobG, CobI and CobJ (aerobic cobalamin biosynthesis), CbiX and CbiL (anaerobic cobalamin biosynthesis), and BluB (synthesis of 5,6-dimethylbenzimidazole, a component of vitamin B12) were among the differentially abundant KO terms in cases.

**Figure 2.**
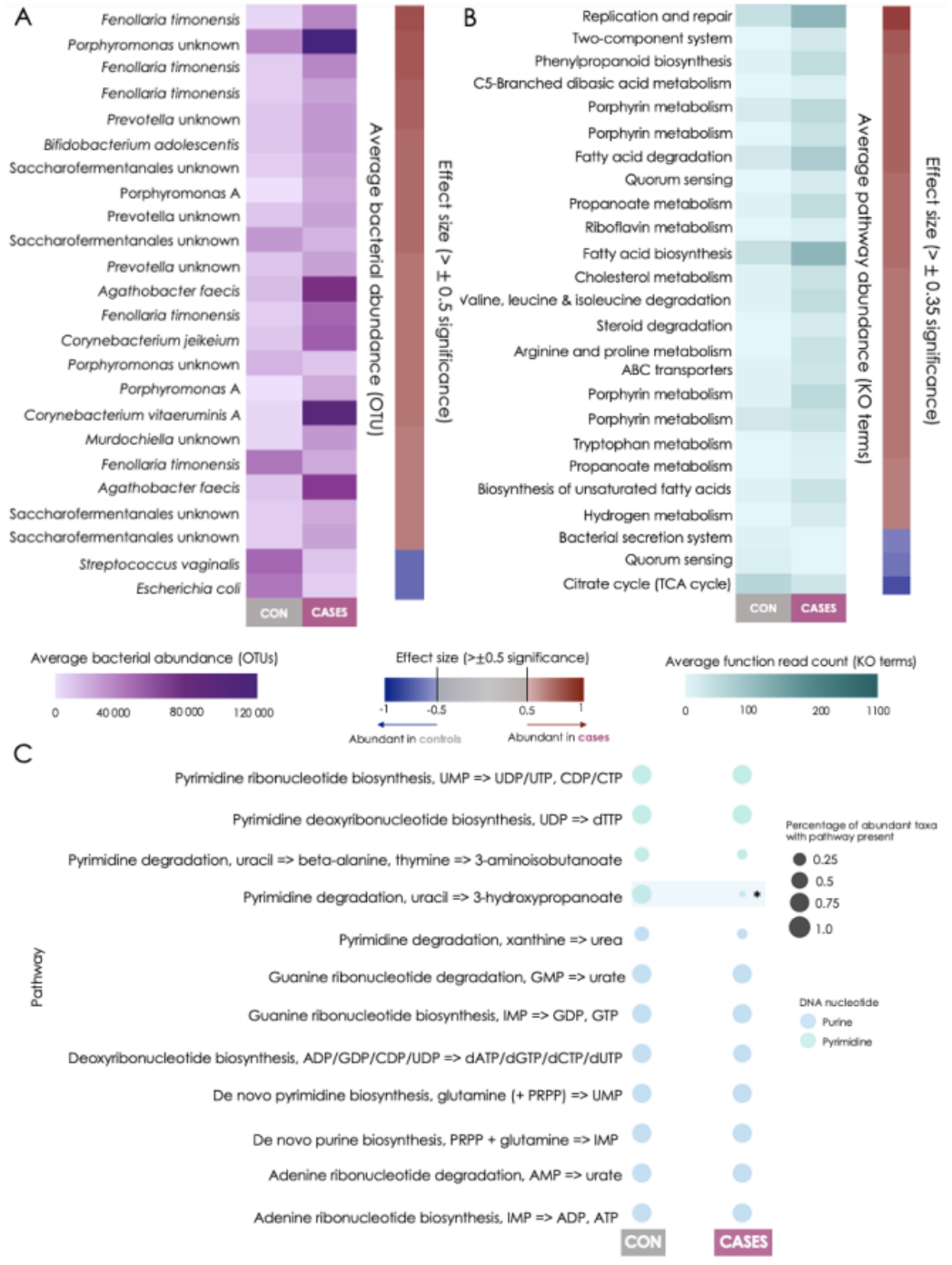
Differentially abundant taxa and functions of the gut microbiome, including presence of microbial DNA biosynthesis pathways in overrepresented taxa. A. Differentially abundant taxa in cases and controls. Differentially abundant taxa in cases are those that are most abundant in cases compared to controls (more overrepresented in cases, effect size > 0.5), and in controls are those that are overrepresented in controls (effect size < -0.5). Average bacterial abundance (OTUs) was plotted in a heatmap to show difference in abundance for each differentially abundant taxon between cases and controls. Effect size heatmap shows the difference in bacterial abundance between groups, where red suggests overrepresentation in cases and blue represents overrepresentation in controls. B. Differentially abundant pathways in cases and controls. Differentially abundant pathways in cases are those that are most abundant in cases compared to controls (more overrepresented in cases, effect size > 0.35), and in controls are those that are most abundant in controls compared to cases (effect size < -30.5). Average pathway abundance was plotted in a heatmap to show difference in abundance for each differentially abundant pathway between cases and controls. Effect size heatmap shows the difference in pathway abundance between groups, where red suggests overrepresentation in cases and blue represents overrepresentation in controls. C. Bubble plots of putative DNA-related KEGG biosynthesis pathways in cases and controls. The size of the bubble represents the percent of differentially abundant taxa with the given pathway and the colour represents the DNA nucleotide/nucleoside involved in the pathways. *Statistically significant, Fisher’s exact test, q<0.05.

Further, lipid and fatty acid metabolism functions were overrepresented in cases compared to controls, including KO terms corresponding to methylmalonyl-CoA carboxyltransferase subunits (propanoate metabolism), DesA3 (oleic acid production), BccA (malonyl-CoA production), and FadH (unsaturated fatty acid beta-oxidation). Finally, functions related to amino acid metabolism were among the more common pathways overrepresented in cases including: HyuB (sarcosine synthesis), AccD (leucine degradation) and tdc (tryptophan degradation). Conversely, only three pathways were overrepresented in controls compared to cases (Figure 2B). Finally, to explore differences in metabolic pathways known to be important in NTDs between overrepresented taxa in cases and controls (d>0.5), pathway counts were aggregated and compared. Fisher’s exact testing revealed a significant relationship between cases and presence of the pyrimidine degradation pathway (q<0.05). There were also fewer pyrimidine degradation functions identified in differentially abundant taxa in cases compared to controls (Figure 2C).

### Gut microbial biosynthetic capability for B vitamins and nucleotides is similar between groups

To determine whether the enrichment of cobalamin biosynthesis functions observed in cases was reflected in greater overall gut microbial B12 biosynthetic capability, and to characterise folate and nucleotide biosynthetic potential more broadly given their roles in development, we estimated individual-level capacity scores for folate, vitamin B12, and purines and pyrimidines. Although complete synthesis pathways were encoded by the majority of taxa (94% of taxa for folate, 59% for vitamin B12, and 98% for purine/pyrimidine, whereas 1% encoded complete purine/pyrimidine degradation pathways), total producing capability was not significantly different in cases compared to controls (Figure 3).

**Figure 3.**
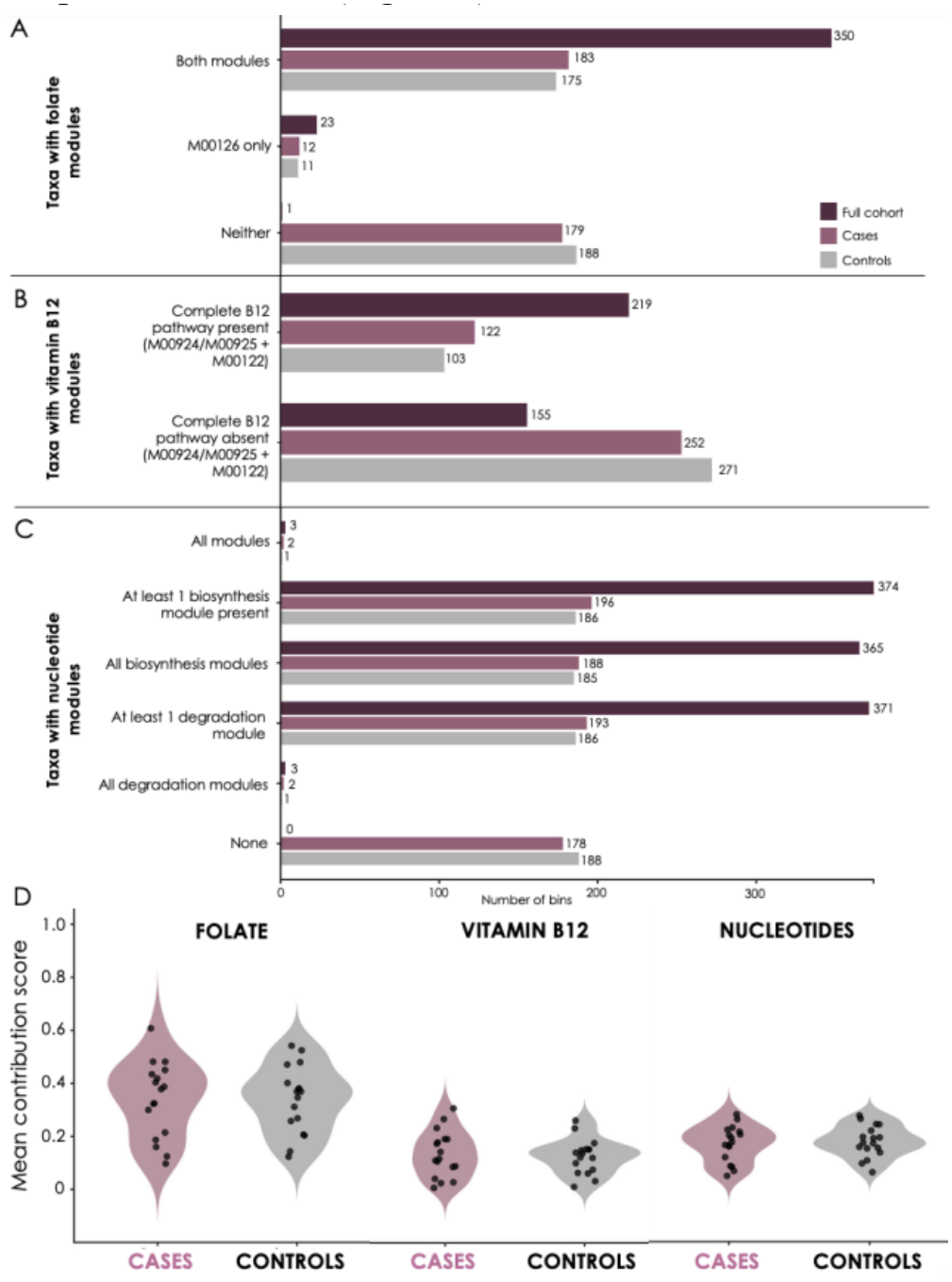
Inferred maternal gut microbial folate, vitamin B12 and DNA nucleotide biosynthetic potential in cases and controls. A. Number of taxonomic binning assignments (species) containing folate biosynthetic KEGG modules M00126 (core de novo tetrahydrofolate biosynthesis from GTP) and M00841 (alternative tetrahydrofolate biosynthesis pathway mediated by para-aminobenzoic acid). B. Number of taxonomic binning assignments containing the complete vitamin B12 biosynthetic pathway, comprising either the anaerobic (M00924) or aerobic (M00925) route for producing cobalamin precursors, together with M00122, which encodes the downstream conversion of cobyrinate a,c-diamide to cobalamin. C. Number of taxonomic binning assignments containing purine biosynthesis (M00048, PRPP to IMP), pyrimidine biosynthesis (M00051, PRPP to UMP), thymidylate biosynthesis (M00938, UDP to dTTP), purine degradation (M00958, AMP to urate; M00959, GMP to urate), and pyrimidine degradation (M00046, uracil/thymine degradation; M00939, uracil to 3-hydroxypropanoate). D. Violin plots representing abundance-weighted inferred microbial biosynthetic contribution scores, where each point represents a participant-level overall contribution score. Cases = pink, controls = grey, and the full cohort = burgundy.

### Circulating metabolite profiles in cases implicate upregulated one carbon and amino acid metabolism with downregulated purine metabolism

To identify differences in maternal circulating metabolites that may reflect altered gut microbial composition and function, we compared individual serum metabolite levels between cases and controls. Cases had significantly higher levels of 28 non-nutrient metabolites compared to controls. These metabolites were mainly amino acids, including alanine, arginine, glutamine, and glycine, among others (Figure 4A). Other key serum metabolites that were higher in cases included lipids and fatty acids (HexCer(d18:1/26:0), LysoPC a C18:2, C14:2), neurotransmitters (3-methoxytyramine, DOPA), and other nitric oxides (ADMA/SDMA) and organic compounds (trimethylamine; Figure 4A).

**Figure 4.**
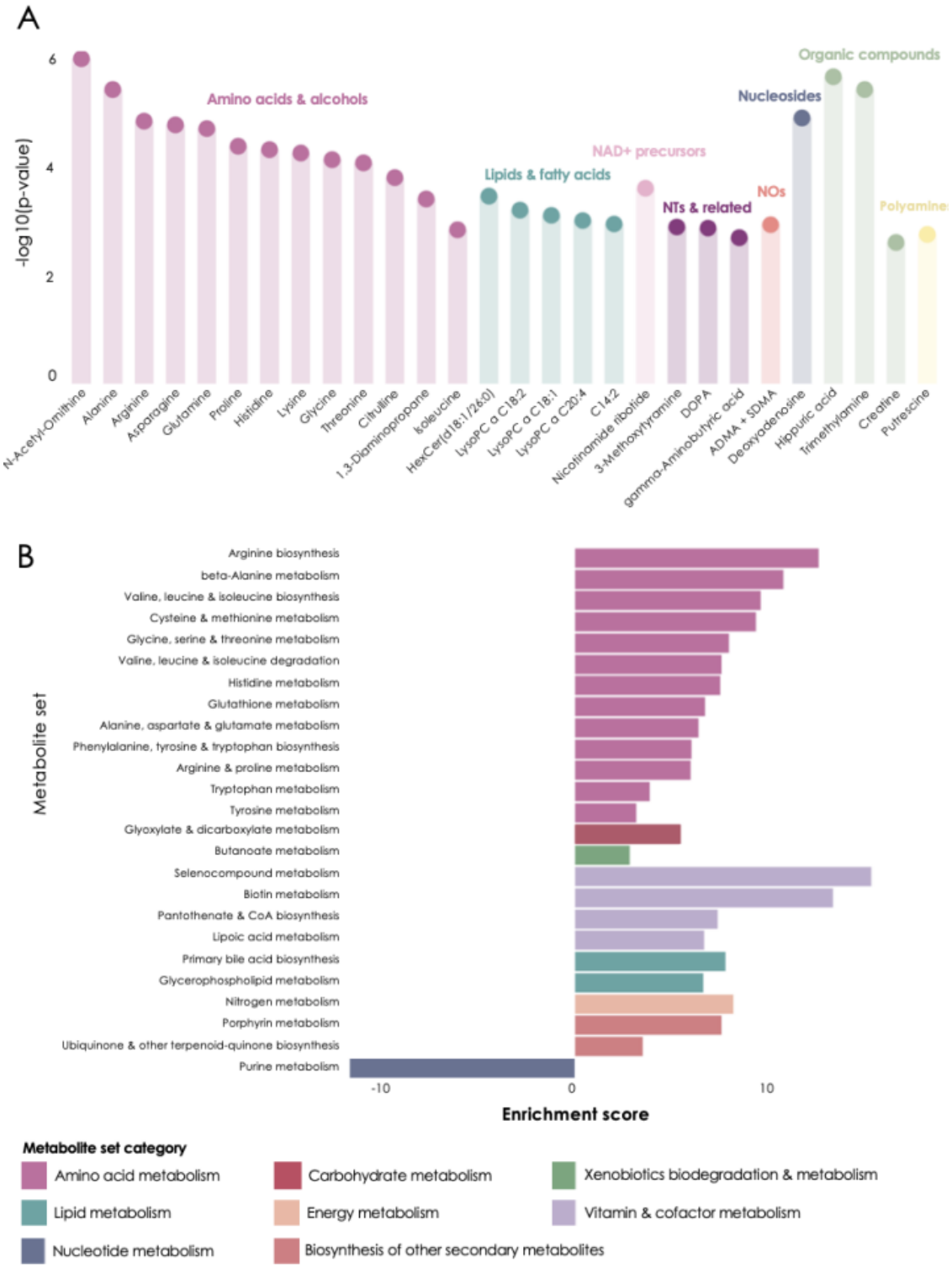
A. Differentially abundant circulating non-nutrient metabolites in cases compared to controls (n=28). Differentially abundant metabolites were determined by Student’s t-test, FDR-corrected, q<0.05 statistical significance. Y-axis represents the level of significance (corresponding to the -log10[p-value]), where metabolites that are higher on the axis have a larger statistical significance. Color of the point on the plot is representing the metabolite category. B. Metabolite set enrichment analysis revealed 25 dysregulated gene sets in cases compared to controls (downregulated in cases: n =1, upregulated in cases: n=24). Dysregulation of metabolite sets (a group of metabolites that share a common biological function or pathway) was determined by computing an enrichment ratio, which is calculated by dividing the number of metabolite sets that are different within a metabolite set by the number of expected different metabolites within that set. Metabolite set enrichment direction (i.e. up or downregulation) was determined by the direction of change of each metabolite within that set. NTs = neurotransmitters; NOs = nitric oxides.

Next, we performed metabolite set enrichment analysis (MSEA), which allows us to group metabolites based on similar metabolic functions, and understand how these metabolic functions may be dysregulated in response to altered metabolite levels. Upregulation of these metabolite sets (enrichment score > 0) suggests increased metabolic activity due to higher metabolite levels, whereas downregulation (enrichment score < 0) suggests decreased metabolic activity due to lower metabolite levels. Consistent with the higher serum levels of the non-nutrient metabolites, we found 24 upregulated metabolite sets in cases compared to controls, mainly related to amino acid metabolism (e.g. arginine biosynthesis, cysteine and methionine metabolism, arginine and proline metabolism), lipid metabolism (primary bile acid metabolism, glycerophospholipid metabolism), and energy metabolism (nitrogen metabolism; Figure 4B), suggesting potential disturbances in metabolic processes involved in nitric oxide production, methylation reactions, and lipid metabolism. One downregulated metabolite set was also identified in cases compared to controls, purine metabolism (Figure 4B), suggesting potentially impaired nucleotide metabolism.

We then performed pathway topological analysis to determine the importance of differentially abundant metabolites based on their position and connections to other metabolites within each pathway. There were 26 dysregulated metabolic pathways in cases compared to controls. Similar to the metabolite set findings, 25 pathways were upregulated, including those related to amino acid metabolism (arginine biosynthesis, alanine, aspartate and glutamate metabolism, tryptophan metabolism), lipid metabolism (primary bile acid biosynthesis, glycerophospholipid metabolism), and vitamin and cofactor metabolism (one carbon pool by folate), and one pathway was downregulated (purine metabolism) in cases (Figure 5).

**Figure 5.**
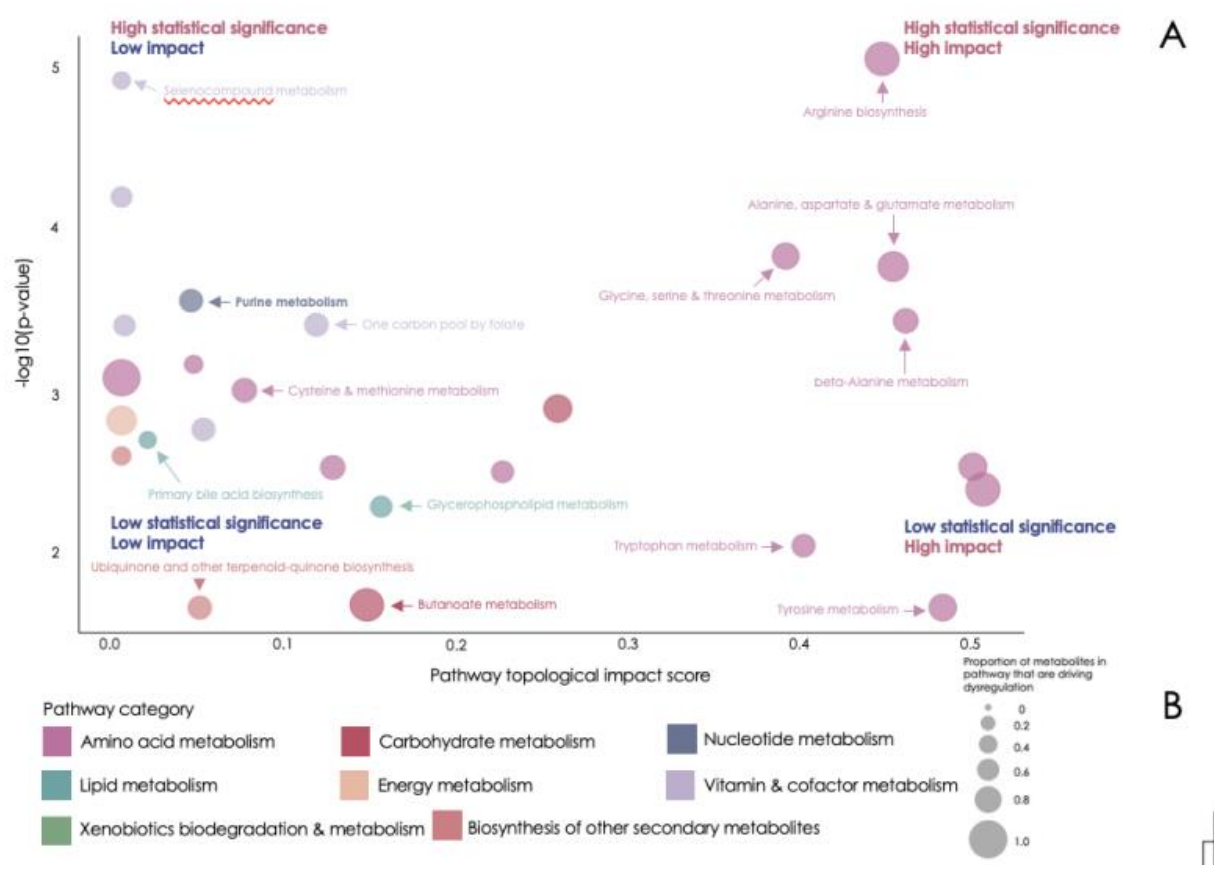
Pathway analysis of 26 dysregulated pathways in cases compared to controls (n=25 upregulated, n=1 downregulated [purine metabolism]). Metabolic pathways were determined by computing pathway impact using topological analysis. This approach considers the position of each metabolite in the network and the number of connections for each metabolite. A higher pathway impact suggests central metabolites within the pathway are changed and the pathway is functionally impacted. .Y-axis represents the level of significance (corresponding to the -log10[p-value]), where metabolites that are higher on the axis have a larger statistical significance. The size of the bubble indicates the proportion of metabolites within the pathway that are driving the dysregulation, and the color indicates the category.

Finally, we performed principal component analysis (PCA) and k-means clustering to visualize overall metabolite distribution (clusters) between cases and controls and identify the metabolites contributing to these clusters. PCA biplots revealed partial separation between cases and controls, primarily along the second principal component (PC2), which explained. 12.5% of the total variance (Figure 6A). The distribution of cases was more closely associated with amino acids, including isoleucine and citrulline, and other metabolites, including indole, whereas the distribution of controls was more closely associated with triglycerides (Figure 6A). To further characterize metabolic variation across the entire cohort, hierarchical clustering of the top 25 metabolites driving clustering of groups was conducted and revealed similar patterns. Metabotype 1 was largely represented by higher concentrations of triglycerides, and lower levels of other metabolites, such as butyric acid, norepinephrine, indole, and isoleucine, among others, whereas metabotype 2 was largely represented by lower levels of triglycerides, and higher levels of other metabolites, including those mentioned above (Figure 6B). However, despite the partial separation observed in the PCA biplot, these metabotypes did not clearly distinguish cases from controls, as both groups were represented within each metabotype.

**Figure 6.**
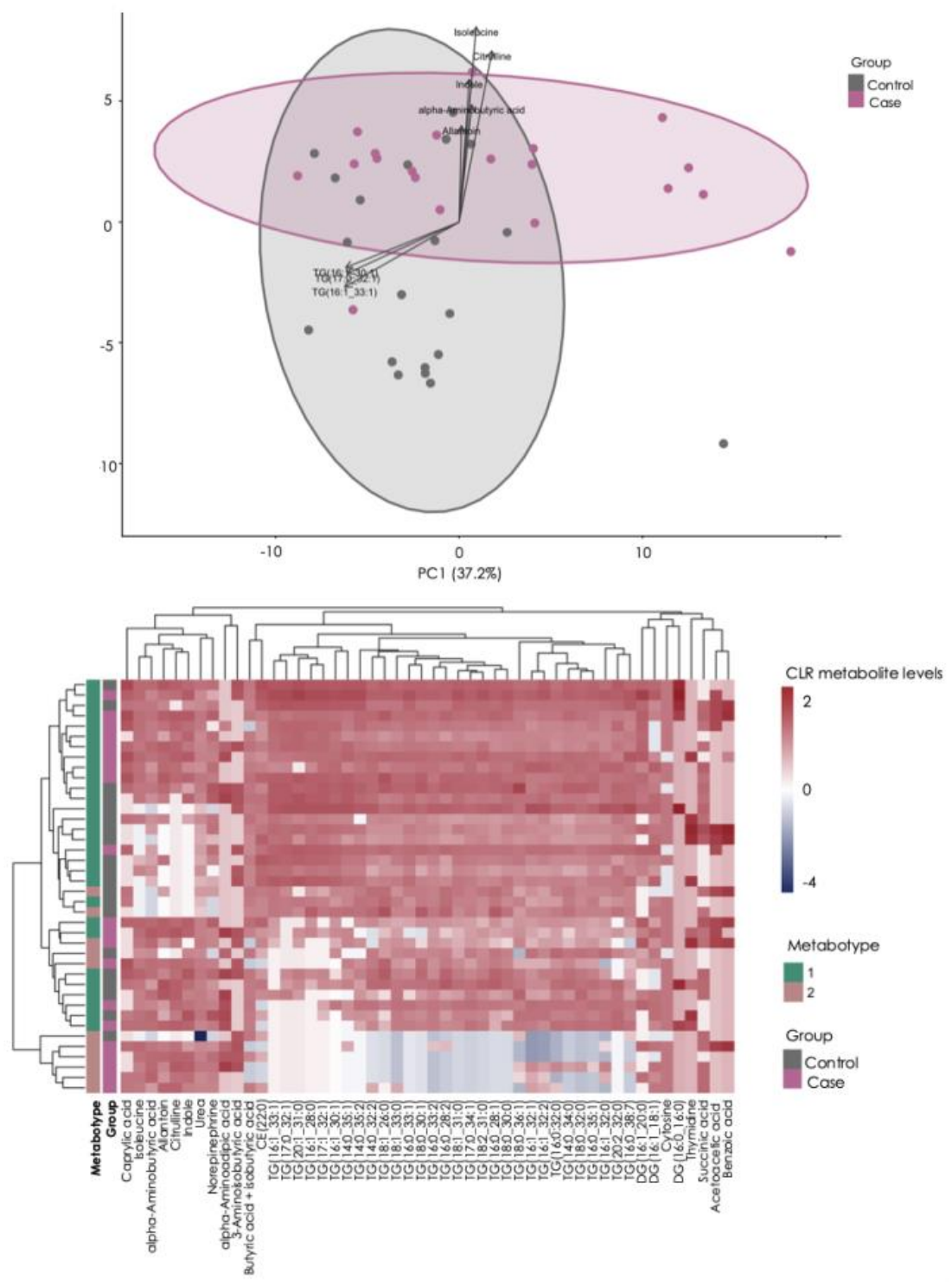
A. Principal components analysis (PCA) biplot of serum metabolites from MEGA assay (amino acids, triglycerides, lipids, neurotransmitters, etc.). Grey dots = controls. Pink dots = cases. B. Clustered heatmap of metabotypes using top 25 metabolites driving clustering (CLR values). Metabotypes are represented by green (metabotype 1) or orange (metabotype 2), and groups are represented by pink (cases) and grey (controls).

### Gut microbial functional profiles covary with circulating amino acids in cases and triglycerides in controls

The maternal gut microbiome may influence circulating metabolite profiles through its metabolic activities. To investigate whether these relationships differed between cases and controls, we integrated our metagenomic and metabolomic data to identify potential links between gut microbial function and maternal circulating metabolites. Specifically, serum levels of all measured metabolites were correlated with the principal components of all samples on the compositional biplots of the KO-aggregated metagenomic data. Correlation coefficients with an absolute value >0.4 were scaled to the sample loadings and overlaid on the plots to visualize relationships between the most influential circulating metabolites and gut microbial functions. For water- and fat-soluble vitamins and metal assays, most metabolites clustered near the origin, with no discernable differences between groups. For metabolites measured in the MEGA assay, multiple triglycerides localised where most of the controls were located (Figure 7).

Conversely, numerous amino acids, their derivatives, and short chain fatty acids were located in close proximity to case samples, including several metabolites such as DOPA, tryptophan, and glutamine, among others, which have previously been associated with NTDs in metabolomic studies^36^.

## Discussion

Despite widespread folic acid fortification and supplementation^12^, NTDs persist^15^, suggesting that factors beyond folate contribute to NTD risk and associated comorbidities. In this case-control study, we identified distinct maternal gut microbial and circulating metabolic signatures in pregnancies affected by fetal spina bifida, despite comparable folate status. Cases had lower serum levels and dietary intake of vitamin B12, along with enrichment of gut microbial cobalamin biosynthesis genes, suggesting a potential compensatory response to reduced vitamin B12 availability. These differences coincided with reduced gut microbial pyrimidine degradation and downregulated circulating purine metabolism, indicating potential disturbances in nucleotide metabolism. Further, differentially abundant gut microbial taxa associated with inflammation and adverse pregnancy outcomes, lower antioxidant nutrient availability (e.g. lower serum vitamin E levels and lower dietary intake of vitamin C), and upregulated circulating amino acid and antioxidant-related metabolism, were consistent with a pro-oxidant maternal metabolic environment. Integration of metagenomic and metabolomic datasets also revealed that microbial amino acid and lipid metabolic functions clustered with circulating metabolites enriched in cases, including amino acids suggesting coordinated alterations in gut microbial metabolic activity and maternal circulating metabolic profiles. Collectively, these findings identify distinct and interconnected gut microbial and systemic metabolic alterations in pregnancies carrying a fetus with spina bifida that extend beyond folate status, raising the possibility that the maternal gut microbiome represents a modifiable contributor to NTD risk and associated comorbidities, and revealing novel targets for periconceptional prevention.

Consistent with the high prevalence of folate sufficiency in this population^13^, folate intake, serum folate levels, and microbial folate-producing potential in cases were similar to controls in our study. This suggests that maternal folate intake, circulating folate status, and gut microbial folate biosynthetic potential may not be the primary factors underlying spina bifida pathogenesis in our folate-replete Canadian cohort. However, we did find alterations in other nutrients within the same metabolic network as folate. Vitamin B12 and choline dietary intakes were lower in cases compared to controls, and circulating vitamin B12 levels were also lower in cases, consistent with our hypothesis. Vitamin B12 and choline supply cofactors and methyl groups for the methionine cycle^37^. Low vitamin B12 level impairs methionine synthase, blocking conversion of 5-methyltetrahydrofolate to the biologically active tetrahydrofolate needed for methylation processes and nucleotide synthesis^38,39^, which are critical for neural tube closure. Thus, a low maternal vitamin B12 level is a known risk factor for NTDs independent of adequate circulating folate levels^11^. Despite lower dietary intake and serum levels of vitamin B12, the gut microbiome of cases was enriched in genes for both aerobic (*cobG, cobI, cobJ*) and anaerobic (*cbiX, cbiL*) cobalamin (B12) biosynthesis. Given most gut microbes require exogenous B12 as a cofactor for essential metabolic processes including DNA synthesis and methylation reactions^9^, the enrichment of cobalamin biosynthesis genes in cases may reflect a compensatory response to reduced vitamin B12 availability within the gut microbial community^40^. These findings suggest that the nutrient-related mechanisms contributing to NTD pathophysiology extend beyond folate alone, particularly in folate-replete populations. Preconception assessment of vitamin B12 levels, followed by targeted vitamin B12 supplementation in women with insufficiency or deficiency, may therefore be a complementary approach to existing folic acid-based prevention strategies for fetal NTDs, warranting evaluation in prospective studies beginning preconceptionally.

Further, we found that purine metabolism was the only circulating metabolic pathway downregulated in cases, and the overrepresented taxa in cases had fewer pyrimidine-degrading functions than controls. This suggests a shared perturbation in nucleotide metabolism across the maternal gut microbiome and maternal circulation, despite no differences in the potential microbial nucleotide biosynthetic capacity in our study. Purine and pyrimidine metabolism are critical for DNA and RNA synthesis and repair^41^, particularly during the rapid cell division occurring during neural tube closure^18,42^ and throughout gestation for optimal fetoplacental growth^43^, and impaired nucleotide metabolism is an established risk factor for NTDs^44^. Lower circulating vitamin B12 levels may also compromise nucleotide synthesis, as insufficient vitamin B12 can impair methionine synthase activity and tetrahydrofolate regeneration^45^, potentially limiting the availability of folate derivatives needed for purine and thymidylate synthesis^38,39^. Collectively, these findings suggest that the changes we observed in the maternal gut microbiome and circulating metabolome in pregnancies with NTDs extend beyond folate, and may reflect disturbances in the one-carbon metabolic processes that depend on vitamin B12 and support nucleotide synthesis.

Oxidative stress has also been implicated in NTD pathogenesis^46^, as high reactive oxygen species (ROS) levels disrupt neural tube closure and impair fetal growth^46^. Novel evidence for a maternal environment permissive of oxidative stress and inflammation was identified across dietary, microbiome, and metabolomic outcomes in our study. Cases reported lower dietary intake of vitamin C and had lower serum vitamin E levels (alpha-tocopherol), both important antioxidants, which is consistent with previous studies linking lower vitamin C intake to higher NTD risk^47,48^. Further, vitamin E deficiency impairs neural tube closure^49^, and vitamin E supplementation can protect against oxidative stress-induced NTDs^50^, highlighting the importance of vitamin E as a protective factor for NTDs. Additionally, whilst total DII scores did not differ between groups, cases had a positive (pro-inflammatory) DII score. The gut microbiome of cases was also enriched with taxa with established links to inflammation and adverse pregnancy outcomes, although their relevance to NTDs has not been previously documented. For example, bacterial species from the *Fenollaria, Porphyromonas*, and *Prevotella* genera were differentially abundant in cases, and greater abundance of species of these genera have been associated with pro-inflammatory conditions and pregnancy complications. Increased abundance of gut *Prevotella* species has been associated with insulin resistance and obesity in non-pregnant people^51^, and greater abundance of gut *Fenollaria*^52^ and oral *Porphyromonas*^34^ species in pregnancy have been implicated in preterm birth. *Porphyromonas* and *Prevotella* are anaerobes that rely on amino acid and peptide metabolism for energy^53,54^, which is consistent with our gut microbial functional findings; amino acid degradation genes (e.g. tyrosine decarboxylase [*tdc*], acetyl-coenzyme A carboxylase carboxyl transferase subunit beta [*accD*]) were overrepresented in cases. Case mothers also had higher serum levels of arginine and asymmetric dimethylarginine (ADMA)/symmetric dimethylarginine (SDMA). These metabolites are mechanistically related to oxidative stress and inflammation. For example, ADMA is an endogenous inhibitor of nitric oxide synthase (NOS), where higher circulating levels block production of protective nitric oxide from arginine^55,56^, leading to endothelial dysfunction and adverse outcomes, such as preeclampsia^57^.

In response to increased oxidative stress, antioxidant defence pathways may be upregulated to maintain redox balance^58^. Consistent with this we observed upregulation of circulating cysteine, ubiquinone, and lipoic acid metabolism case mothers. Previous studies have identified higher circulating levels of glutathione (an intracellular antioxidant) and cysteine (required for glutathione synthesis) in mothers carrying a fetus with NTDs^59^, consistent with upregulated cysteine metabolism in our study. Importantly, this interpretation is supported by our previous finding of upregulated serum glycerophospholipid metabolism, which is consistent with previous evidence showing upregulated fetal brain glycerophospholipid metabolism in a human study and mouse model of NTDs^69^. Phospholipid synthesis is closely linked to one-carbon metabolism, as vitamin B12 supports the generation of methyl donors required for phosphatidylcholine synthesis through the phosphatidylethanolamine N-methyltransferase (PEMT) pathway, while choline serves as a direct precursor for phosphatidylcholine synthesis^70^. Thus, the lower dietary choline intake and circulating vitamin B12 levels observed in cases may be linked to altered glycerophospholipid metabolism, suggesting that disturbances in one-carbon and upregulated placental catalase (*CAT*; a placental gene responsive to oxidative stress) in this same cohort^5^. *<u>CAT</u>* was amongst the most differentially expressed placental genes in spina bifida. The cooccurrence of elevated antioxidant-responsive metabolic pathways in maternal circulation and upregulated antioxidant enzyme expression in placental tissue across the same pregnancies provides cross-tissue evidence of a sustained oxidatively stressed pregnancy environment with spina bifida.

Whilst samples were collected at 25 weeks’ gestation, the persistence of this pro-oxidant signature into mid gestation has implications beyond the initial failure of neural tube closure. This includes contributing to placental dysfunction^60^, poor fetal growth^61^, and preterm birth risk^62^, which are known comorbidities of NTDs^5,6^. Importantly, the two-hit hypothesis of spina bifida proposes that prolonged exposure of the unprotected spinal cord to the intrauterine environment results in progressive neural tissue damage throughout gestation^63^. Recent evidence of altered oxidative-antioxidative balance in the cerebrospinal fluid of fetuses with spina bifida suggests that oxidative stress may contribute to this secondary tissue injury^64^. This may also have implications for fetal surgical repair, as excessive oxidative stress can impair wound healing by promoting inflammation and disrupting tissue regeneration^65^. The alterations in oxidative-stress related metabolism and antioxidant defence pathways identified in our study therefore point to maternal redox regulation as a potential target for NTD prevention and improving fetal outcomes in these pregnancies.

Lipids are essential substrates for fetal growth and placental function^66^, and the gut microbiome is a key regulator of host lipid metabolism^67^. Here we found that cases had enriched gut microbial pathways related to lipid metabolism (e.g. cholesterol metabolism) and fatty acid metabolism (e.g. biosynthesis of unsaturated fatty acids), including *DesA3, BccA*, and *FadH*, and had higher serum levels of fatty acids and lipids (e.g. LysoPC a C18:3, LysoPC a C20:4). Interestingly, gut microbial pathways related to fatty acid and lipid metabolism were enriched in a previous study of pregnant people with higher risk of gestational diabetes, a known risk factor for NTDs^68^. We also found that cases had lipid metabolism may be interconnected in pregnancies affected by fetal spina bifida. Circulating short-chain fatty acids, which clustered with case samples in our serum metabolomics data, regulate host lipid and glucose metabolism and inflammation^71^. Further, maternal obesity and diabetes are established risk factors for NTDs^68^, both of which are conditions of disordered lipid and glucose handling^72^, and the gut microbiome has been implicated in their pathophysiology through its role in regulating lipid and fatty acid metabolism^67^, and production of potentially harmful metabolites, such as trimethylamine (TMA)^73^. Interestingly, in our study, cases had higher serum levels of TMA, and higher circulating levels of TMA have been previously associated with gestational diabetes^73^. In our study, because groups were matched for BMI, dysregulated lipid metabolism and increased lipid-related metabolite levels in cases are unlikely to reflect classical metabolic diseases, such as obesity and diabetes. They may instead reflect altered maternal lipid handling to increase lipid availability to support a sub-optimally growing fetus, consistent with poor fetal growth seen with NTDs^6^, or to support greater placental demand for lipid substrates, as there is evidence of suboptimal placental growth and function in pregnancies with NTDs^74^. Taken together, the enrichment of gut microbial lipid and fatty acid metabolism pathways, higher circulating lipid and fatty acid levels, and upregulation of serum glycerophospholipid metabolism in cases suggest that maternal lipid metabolism may be altered in pregnancies affected by fetal spina bifida, with potential implications for fetoplacental growth.

A major strength of our study is the multi-omics design. The maternal gut microbiome synthesizes and degrades substrates required for neural tube closure and fetoplacental development, including B vitamins and nucleotides^16,17,75,76^, among others. Because fetuses with spina bifida have known comorbidities, including poor growth^6^, it is imperative to investigate whether an altered maternal gut microbiome contributes to altered availability of substrates important for developmental processes like neural tube closure. Using shotgun metagenomic sequencing, we were able to obtain greater taxonomic and functional resolution compared to other traditional methods^77^, allowing us to fully capture the biosynthetic and metabolic capabilities of the gut microbiome. By analyzing circulating metabolomics data, and integrating this with our metagenomic data, we were also able to determine if alterations in the gut microbiome were reflected in maternal circulation, providing additional insights into potential metabolic disruptions, including in pathways related to one-carbon metabolism, associated with NTDs and their comorbidities. However, there are some limitations. Our samples were collected at approximately 25 weeks’ gestation, after neural tube closure, which occurs around four weeks post-conception^78^. Our data therefore characterize the maternal gut microbial and metabolic environment at mid-gestation, not during the period in which the defect occurred. However, longitudinal studies have demonstrated that the composition of maternal gut microbial communities remains relatively consistent across pregnancy, although the relative abundances of individual bacterial taxa may change over time^32^. Similarly, longitudinal metabolomic studies have shown that while circulating metabolites, including amino acids and fatty acids, exhibit varying trajectories throughout gestation, many remain stable between early and mid-gestation^79^. Thus, differences in gut microbial composition and functional capacity observed at mid-gestation may reflect maternal states that were present during the early pregnancy period. Nevertheless, longitudinal studies, including those beginning preconception, are required to establish the temporal relationships between the maternal gut microbiome and metabolic differences, and to determine whether these relate to fetal growth and placental outcomes in pregnancies carrying a fetus with spina bifida. Another limitation is our sample size, which reflects the low prevalence of NTDs in populations where folate fortification and preconception folic acid supplementation are standard^80^, and thus, recruiting a larger cohort is not feasible. However, our study met the minimum sample size established by our *a priori* power calculation, and cases and controls were matched for maternal age, BMI and gestational age at recruitment, thereby limiting potential confounding by these factors. Nonetheless, our study is one of the first multi-omics investigations of the human maternal gut microbiome and circulating metabolome in pregnancies affected by fetal spina bifida, providing novel insights into the multifactorial nature of NTD pathophysiology.

Our multi-omic findings provide a new reframing of the maternal metabolic context of fetal spina bifida. Beyond folate, our data identify vitamin B12 insufficiency, disrupted nucleotide metabolism and a pro-oxidant gestational environment, present across the maternal gut microbiome and circulating metabolome, as likely interconnected contributors to risk for NTDs and their comorbidities. The consistency of these signatures across multiple biological sites strengthens the case that NTD pathophysiology extends beyond folate status, and positions the maternal gut microbiome as a previously underappreciated link between nutritional status and developmental outcomes. Longitudinal studies beginning preconceptionally are needed to establish whether these metabolic signatures precede NTD occurrence, and if confirmed, would support expanding periconceptional screening to include other micronutrients including vitamin B12, and open avenues for microbiome-targeted nutritional interventions as complements to folic acid supplementation to improve maternal periconceptional and fetal health.

## Data Availability

All data produced in the present study are available upon reasonable request to the authors.

## Acknowledgments

This research is supported by the Canadian Institutes of Health Research (CIHR PJT-175161), and research development grants from the Department of Obstetrics and Gynecology at Mount Sinai Hospital, Toronto, Canada and Carleton University, Ottawa, Canada. HS was supported by an Ontario Graduate Scholarship, Faculty of Science, Carleton University.

## Author Contributions and Notes

Conceptualization: HS, SDS, GBG, TVM, KLC; Methodology: HS, SDS, GBG, TVM, KLC; Funding acquisition: TVM, KLC; Resources: VP, AJM, TVM, KLC; Investigation: HS, SDS, BRJ, VP, AJM, KLC; Data curation: HS, SDS, BRJ, VP, AJM; Formal analysis: HS, SDS, BRJ;

Software: GBG; Validation: HS, SDS, GBG, KLC; Visualization: HS, SDS, KLC; Project administration: HS, VP, TVM, KLC; Supervision: GBG, TVM, KLC; Writing – original draft: HS, SDS, KLC; Writing – review & editing: HS, SDS, VP, AJM, GBG, TVM, KLC.

## Supplementary methods

### Bacterial DNA extraction and analyses

Total genomic DNA was isolated from rectal swabs of cases (n=17) and controls (n=17) using the QiAmp Powerfecal Pro DNA Kit, per manufacturer’s protocol and sequenced on an Illumina NovaSeq to produce 75 nt paired end reads (London Regional Genomics Centre, London, ON).

### Assembly and binning of metagenomes

Raw reads were mapped against two human genomes using Bowtie2^1^: the GRCh38 assembly and the complete, telomere-to-telomere assembly chm13v2.0^2^, retaining only reads that did not align; these are referred to as non-human reads. Non-human reads were de-duplicated using the BBMap package and concordant read pairs were extracted from the dereplicated FASTQ files. FastQC^3^ and MultiQC^4^ were used to assess the quality of dereplicated, non-human reads and identify a suitable quality threshold for trimming reads.

Based on the multiQC report, reads were trimmed to 74 nt with Trimmomatic^5^ prior to assembly of metagenomic contigs using a combination of metaSPAdes^6^ and MEGAHIT^7^, implemented in the MetaWRAP^8^ package. Any contigs classified as human by Kraken2^9^ were removed prior to binning with MetaBAT2^10^ and Maxbin2^11^. The resultant bins were refined and reassembled using the respective modules in the MetaWRAP package, retaining only those >75% complete and containing <10% contamination (i.e. at least medium quality bins). Bins were dereplicated at a clustering threshold on 99% similarity using dRep^12^.

### Taxonomic classification and quantification of dereplicated bins

Taxonomy was assigned to dereplicated bins using the Genome Taxonomy Database (GTDB) toolkit^13^ and GTDB release 220. For quantification of bins in the dataset, quality-trimmed, non-human reads were mapped to the dereplicated bins using Bowtie2 and the resultant SAM files were used to construct a feature table of read counts per bin from coverage information generated by samtools^14^.

### Functional classification and quantification within dereplicated bins

Dereplicated bins were concatenated into a single FASTA file and used to construct an Anvi’o contig database for gene-calling with Prodigal^15^. KEGG orthology terms were assigned to ORFs based on hidden Markov model hits (hmmsearch^16^) against the latest version of the KEGG KOfams database via an implementation in the Anvi’o package^17^. Gene names, symbols and pathway memberships were assigned based on the KEGG KO hierarchy^18^ downloaded on 19 December 2024 from: https://rest.kegg.jp/get/br:ko00001. This list of 27,306 KEGG orthology (KO) terms was manually curated to match each KO term with a single KEGG pathway; the resulting KO-pathway lookup table is available on GitHub (https://github.com/scottdossantos/sb_metagenome2025).

Human-depleted, quality trimmed reads from each sample were mapped against the FASTA file of genes called by prodigal and exported from the KEGG-mapped Anvi’o contigs database. The resulting SAM files were used to construct a feature table of read counts per gene from samtools coverage information. This table was then aggregated by KO terms to construct the functional feature table.

### Differential abundance analysis

Prior to any formal analysis, read count tables for taxonomic bins and KEGG functions were batch-corrected in R (v4.3.1) using the ‘ComBat_seq’ function from the sva R package^19^ (v3.5.0). This function models the count data using a negative binomial distribution and estimates the effect of each batch on the count data in terms of the effect on each feature’s mean count and dispersion within the data set. The tool also preserves the effect of the biological conditions on the data (i.e. cases vs. controls). The adjustment subtracts the estimated batch effect from the log of the expected mean counts of the data which were originally modelled at the start, providing a table of expected counts that reflects sequencing data that would have been observed without any batch differences. Differentially abundant taxa and functions (KO terms) were identified using the ALDEx2 R package^20^ (v1.35.0) in R using scale simulation random variables^21,22^. In brief, feature tables of reads per bin or per function were used to build a Bayesian posterior model of the count data. Posterior values included a scaled log-ratio transformation over 128 Monte-Carlo instances, incorporating scale uncertainty of 0.5 standard deviations and a scale difference of 2.5% between case and control groups. This value was empirically identified as the difference necessary to centre housekeeping functions around the point of no-difference-an approach which has been shown to improve normalisation of high-throughput sequencing data^23,24^. ALDEx2 effect sizes were computed as a measure of the magnitude of difference between cases and controls when considering within and between group differences (based on a Wilcoxon Rank sum test, Welch’s t-test, and Kruskal-Wallis test). Bins and functions with absolute effect sizes >0.5 and >0.35, respectively, were considered significant. These cutoffs correspond to a medium effect size (Cohen’s d). ALDEx2 calculates effect sizes that are roughly 70% the size of Cohen’s d^20^. A negative effect size indicates overrepresentation of bins and functions in controls, whereas a positive effect size indicates overrepresentation of bins and functions in cases.

### Exploratory analysis of microbial folate-, vitamin B12-, nucleotide-producing capability

An exploratory analysis was performed to determine the putative folate, vitamin B12 and DNA nucleotide producing capability of each sample in cases and controls. For folate, two key functional modules, M00126 and M00841, were used to determine whether a taxonomic binning assignment could produce folate. The M00126 module, which is the tetrahydrofolate biosynthesis GTP =>THF pathway, and the M00841 module, which is the tetrahydrofolate biosynthesis, mediated by PTPS, GTP=>THF pathway, individually have different folate-producing capabilities^18^. The M00126 module represents the core de novo pathway for tetrahydrofolate synthesis, whereas M00841 represents an alternative route for tetrahydrofolate biosynthesis via p-aminobenzoate (pABA)^18^. A species that has both pathways is likely able to produce folate, depending on whether it has all the necessary pathway components present^25^. For vitamin B12, three key functional modules, M00924, M00925 and M00122, were used to determine whether a taxonomic binning assignment could produce vitamin B12. The M00924 module, which is the anaerobic uroporphyrinogen III => sirohydrochlorin => cobyrinate a,c-diamide pathway, and the M00925, which is the aerobic uroporphyrinogen III => precorrin => cobyrinate a,c-diamide pathway, are both upstream pathways for the production of vitamin B12 precursor molecules^26^. These molecules are then used to produce cobalamin, via the M00122 module, which is the cobyrinate a,c-diamide => cobalamin pathway^18^. Finally, for DNA nucleotides, several modules were identified for nucleotide biosynthesis (M00048, M00051, M00938) and degradation (M00958, M00959, M00046, M00939), and were used to determine the overall nucleotide production capability of the taxonomic binning assignments^18^.

First, using the Anvi’o package, a pathwise module completeness score was computed for all folate, vitamin B12 and purine/pyrimidine modules on a scale of 0-1, where any score above 0.75 was considered a complete pathway^27^. The pathwise module completeness score is a measure of the defined set of enzymatic steps within a pathway, where a higher score indicates more steps present within that pathway. For folate and DNA nucleotide modules, the individual module pathwise completeness scores in each taxonomic binning assignment in each sample was retained for subsequent steps of the analyses. For the vitamin B12 modules, of the two upstream pathways (M00924/M00925), the pathwise module completeness scores were compared in each taxonomic binning assignment within each sample, and the higher of the two scores was selected. This was done to determine which of the two pathways a given microbe is most likely using to produce the B12 precursor molecules (anaerobically or aerobically). Following this, the pathwise module completeness score for M00122 for each taxonomic binning assignment within each sample was compared to the upstream pathway (M00924 or M00925) pathwise module completeness score, to determine the rate-limiting step in B12 production. Whichever completeness score was lower among M00122 or M00924/M00925 was retained as the overall pathwise completeness score for each taxonomic binning assignment within each sample. Next, the completeness scores were multiplied by the relative abundance of the taxonomic binning assignment for each individual sample, to yield an abundance-weighted contribution score for each module individually per taxonomic binning assignment per sample. Third, the abundance-weighted contribution scores for each pathway separately were then summed across all taxonomic binning assignments within a single sample to create: two taxonomic binning assignment-specific folate-producing capability scores, one single taxonomic binning assignment-specific vitamin B12-producing capability score, and seven taxonomic binning assignment-specific nucleotide-producing or degrading capability scores. For folate, the two individual scores were summed to yield a single folate-producing score, whereas for the DNA nucleotides, the biosynthesis scores (M00048, M00051, M00938) were individually summed, the degradation scores (M00958, M00959, M00046, M00939) were individually summed, and the net score was computed (biosynthesis score - degradation score) to determine overall nucleotide production capability. Finally, differences between cases and controls in folate-, vitamin B12- and nucleotide-producing capacity were determined by comparing the mean scores between groups (student’s t-test, FDR *q*<0.05).

### Dietary recall analysis

Dietary recall questionnaires (Automated Self-Administered 24-hour Recall, specific to the Canadian population [ASA24-Canada-2016 & 2018]) were completed at recruitment for cases and controls. Dietary intake data were collected for 18 participants in each group (n=36) The ASA24-Canada-2016 and 2018 are an adapted version of the National Institute of Health’s ASA24, which was developed alongside Health Canada. The ASA24-Canada-2016 and 2018 include data from the Canadian Nutrient File (2015), which is a database that contains nutrient levels for nearly 6000 foods that are commonly consumed by Canadians. The questionnaire was administered through an online platform that allows participants to enter a one-time retrospective dietary recall at their study visit. Output datasets that contain food items and macro- and micronutrient intakes for whole foods and supplements were extracted from the database. Quality control was conducted to remove any outlier entries and identify potential errors, where dietary recall entries with overall kcal intake below 600 kcal/day or above 4400 kcal/day were removed from the analyses, as per guidance provided by the ASA24 platform^28^.

Healthy Eating Index (HEI)^29^ scores were calculated to assess how well maternal dietary intakes matched recommendations from established dietary guidelines in Canada. Scores were calculated using the HEI scoring algorithm method, which includes volume of reported food intake for different food categories that are used to determine the HEI adequacy component scores, moderation component scores, and a total HEI score. The HEI adequacy component scores were calculated for the following food categories: total fruits, whole fruits, total vegetables, greens and beans, whole grains, dairy, total protein foods, seafood and plant proteins, and fatty acids. The HEI moderation component scores represent dietary components that should be limited or consumed in moderation, and were calculated for the following food categories: refined grains, sodium, added sugars, and saturated fats. The total HEI score is the sum of all component scores and was used to classify the diet as “good” (total score > 80), “needs improvement” (total score 51-80), or “poor” (total score < 51).

Dietary inflammatory index (DII)^30^ scores were also calculated to estimate the inflammatory potential of the reported dietary intakes. The scores were calculated using 29 food parameters; 27 nutrients and two food items (onions and garlic). Scores can range from -8.87 to 7.98, where a negative DII score corresponds to a diet with more anti-inflammatory potential, a positive DII score corresponds to a diet with more pro-inflammatory potential, and a score of 0 represents a neutral inflammatory diet.

